# Radial ESWT combined with a specific rehabilitation program (rESWT + RP) is more effective than sham rESWT + RP for acute hamstring muscle complex injury type 3b: a randomized, controlled trial

**DOI:** 10.1101/2025.01.03.24319763

**Authors:** Javier Crupnik, Santiago Silveti, Natalia Wajnstein, Alejandro Rolon, Tobias Wuerfel, Peter Stiller, Antoni Morral, John P. Furia, Nicola Maffulli, Christoph Schmitz

## Abstract

**Objectives:** This study tested the hypothesis that radial extracorporeal shock wave therapy combined with a specific rehabilitation program (rESWT + RP) is more effective than sham rESWT + RP in athletes with acute hamstring muscle complex (HMC) injury type 3b.

**Methods:** This was a prospective, randomized, sham-controlled, single center trial with published protocol, concealed allocation, blinded patients and blinded assessors. A total of 36 semi-professional athletes (soccer, field hockey and rugby players) receiving fees or university scholarships with acute HMC injury type 3b were randomly allocated to rESWT + RP for up to 5 weeks (n=18) or sham rESWT + RP (n=18). The primary outcome was the individual time to return to sport. Secondary outcomes were the individual patient’s satisfaction and presence or absence of re-injury during 6 months post-inclusion into this trial.

**Results:** No serious adverse events occurred during the trial. The median / mean time to return to sport was 25.5 / 25.4 ± 3.5 (mean ± SD) days after rESWT + RP (n=18) and 27.5 / 28.3 ± 4.5 days after sham rESWT + RP (n=18) (p=0.037). The mean patient’s satisfaction was not significantly different between the groups. Only one patient in each group experienced a re-injury during 6 months post-inclusion into this trial.

**Conclusion:** In rehabilitation of athletes with acute HMC injury type 3b, rESWT + RP is more effective than sham rESWT + RP.

**Registration:** ClinicalTrials.gov ID NCT03473899 (registered on March 22, 2018).

## INTRODUCTION

Acute hamstring muscle complex (HMC) injuries are among the most common sports injuries in general and are the most common of all sports-related injuries in soccer [1]. Recurrent HMC injuries are common at up to 25% recurrence rate within one year [2].

According to the classification of the Munich Consensus Statement [3] muscle injuries are divided into non-structural (types 1 and 2) and structural (types 3 and 4) injuries. In this classification, the structural muscle injuries are further divided into partial lesions of minor extent (<5 mm, type 3a), partial lesions of greater extent (>5 mm, type 3b) and complete muscle tears or tendinous avulsion (type 4) [3].

Treatment can be challenging. While type 4 injuries often require surgical treatment [4], partial lesions (type 3a and 3b) are usually treated conservatively. The most promising treatment modality for a partial muscle lesion (type 3a and 3b) is a progressive physical therapy training program [5-7]. Other options, such as injection therapy with biologic agents, have had mixed results (e.g., [8]).

Recovery from type 3a and 3b injuries with traditional physical therapy can be lengthy and take up to six weeks to return to sport [3]. Furthermore, the elevated risk of re-injury following acute HMC injuries underscores the potential inadequacy of commonly employed rehabilitation programs. Consequently, there is a demand for the development of innovative treatment approaches, especially for acute HMC injury type 3b.

Over the last decades, extracorporeal shock wave therapy (ESWT) has become established as an effective and safe non-invasive treatment option for various pathologies of the musculoskeletal system [9-11]. Basic science studies and investigations of animal models have suggested that ESWT may accelerate the recovery of muscle injuries [12-14], which has been confirmed in case reports and case series [15,16]. On the other hand, randomized controlled trials (RCTs) testing the efficacy and safety of ESWT for acute HMC injury type 3b have not yet been published.

This study tested the hypothesis that the combination of radial ESWT (rESWT) and a specific rehabilitation program (RP) is effective and safe in treatment of acute HMC injury type 3b in athletes, and is more effective than the combination of sham-rESWT and RP. Because of the proven efficacy and safety of rESWT in the treatment of chronic proximal hamstring tendinopathy in professional athletes [17] rESWT was also used in this study.

## METHODS

### Study Design

This trial was a randomized, controlled, superiority trial in which patients were randomized to either rESWT + RP or sham rESWT + RP. The assessors (SS and NW) and the statistician (CS) were blinded. The protocol was published [18] and the trial was prospectively registered on March 22, 2018 (ClinicalTrials.gov ID NCT03473899). All patients (semi-professional athletes receiving fees or university scholarships) were recruited from a sports club in the Province Buenos Aires (Argentina) between May 2018 and December 2020. Officials of the club were instructed that athletes who experience sudden, sharp pain in the posterior aspect of the thigh during training or competition shall immediately stop activity and come to the clinic of the Principal Investigator (JC) (hereafter: PI). These athletes were then evaluated regarding the presence of the inclusion criteria of this trial. Patients were assessed at baseline, on each day when rESWT or sham rESWT was performed (interval between treatments: two or three days), and afterwards on every second day until return to sport was achieved.

The randomization scheme was generated by a medical assistant at the clinic of the PI with the use of the website, www.randomization.com and was not disclosed to anyone. Forty patients were randomized into five blocks. Interventions were allocated by means of opaque, sealed envelopes that were marked according to the allocation schedule. The randomized intervention assignment was concealed from both patients and assessors until recruitment was complete and irrevocable. Once a patient completed the baseline assessment, the PI opened the next opaque sealed envelope and retrieved the patient’s group allocation. The patient was deemed to have entered the trial at this point. No patient was replaced after randomization.

The PI who administered rESWT or sham rESWT was not blinded, as experienced rESWT therapists can determine whether the treatment is real, or a sham based on the patient’s response to the treatment (according to its working principle an effective rESWT treatment must hurt to some extent). The solution to this issue was a strict, standardized way of interaction between the PI and the patients, irrespective of treatment allocation (c.f. [19, 20]).

### Patients, Therapists, Centers

Adults aged 18-35 years (both female and male) with clinical and ultrasonographic diagnosis of acute HMC injury type 3b were eligible for inclusion.

The inclusion criteria were: physical conditions for rehabilitation (i.e., no surgery required), willingness of the patient to participate in the trial, written informed consent signed and personally dated by the patient, and no contraindications for rESWT.

The exclusion criteria were: children and teenagers below the age of 18, adults aged >35 years old, patients with clinical and ultrasonographic diagnosis of acute HMC injury type 3b who got injured more than seven days before potential enrollment into this trial, patients with clinical and ultrasonographic diagnosis of acute HMC injury type 3a or type 4, bilateral acute HMC injury (types 3a, 3b or 4), proven or suspected HMC injury (types 3a, 3b or 4) of the same lower limb in the time period of six months before potential enrollment into this trial, muscle injury caused by external impact on the posterior aspect of the affected thigh (Category B according to [3]), surgery on the affected lower limb within one year before potential enrollment into this trial, acute or chronic lumbar pathology (as some cases of thigh pain may relate to spinal pathology; c.f. [21]), no willingness of the patient to participate in this trial, written informed consent not signed and not personally dated by the patient, and contraindications to rESWT (including treatment of pregnant patients, patients with blood-clotting disorders (including local thrombosis), patients treated with oral anticoagulants, patients with local bacterial and/or viral infections/ inflammations, patients with local tumors, and patients treated with local corticosteroid applications in the time period of six weeks before the first rESWT session (if applicable)).

### Intervention

All patients performed a specific rehabilitation program (RP) that lasted for eight weeks, independent of the individual time to return to sport (in line with [22]). This RP was developed based on recommendations in the literature [23-25]). The key objective of this RP was that after injury, the patient developed functional, neuromuscular and biomechanical skills according to the demands of the sport she/he performed, while minimizing the risk of re-injury. Therefore, the developed RP guided the patient through a combination of low-risk and high-demand movements, based on a systematic process. This process consisted of an orderly sequence of steps or phases – acute phase, subacute/ regeneration phase, and functional phase. Each phase depended on the outcome of the previous phase and used the individualized response as criterion of progression. The RP was controlled by a co-worker (SS) at the clinic of the PI; this co-worker did not participate in the inclusion/exclusion process at baseline. The PI did not participate in any subsequent evaluation of the patient.

The goals of the acute phase (Phase I) included to (i) prevent re-rupture at the injured site, (ii) prevent excessive inflammation and formation of scar tissue, (iii) increase tensile strength, adhesion and elasticity of new granulation tissue, (iv) reduce build-up of interstitial fluid, and (v) detect and treat any lumbo-pelvic dysfunction. Once a patient was included in the trial, she/he was instructed to avoid the use of medication and apply the RICE principle (rest, ice, compression and elevation) to stop the injury-induced bleeding into the muscle tissue and thereby minimize the extent of the injury (c.f. [26]).

The criterion for progression to the subacute/regeneration phase was absence of pain, which was achieved after 3 to 4 days. No patient in the trial required special attention and treatment by an orthopedic surgeon for the presence of more extensive tissue damage and/or intramuscular hematoma, indicated by persisting muscle pain for more than five days according to the protocol.

The goals of the subacute/regeneration phase (Phase II) included to (i) improve overall core stability, (ii) improve strength and symmetry, (iii) reduce pain during prone isometric, isolated hamstring contractions at 15° knee flexion, (iv) improve hamstring flexibility of both legs, (v) improve hip flexor flexibility of both legs, and (vi) improve neuromuscular control. During the subacute/regeneration phase the patient worked on both legs daily during a single session. On the days when rESWT or sham rESWT was applied, these exercises were performed at the clinic of the PI; on the other days, these exercises were performed at the club. Exercises were conducted to correct the different risk factors and mechanisms related to the lesion of the hamstring musculature. The exercises were divided into four groups: core stability and lumbopelvic control, flexibility and neural mobilization, hamstring and gluteal strength, and running technique. In addition, basic aerobic conditioning started when the patient was able to perform at least three sessions of running without any discomfort or pain. Three running sessions per week were performed at the clinic of the PI and included four sets of five minutes at a low to moderate intensity (individually rated by the patient). Suspension of running sessions was permitted in the event of discomfort or pain.

The criteria for progression to the functional phase were: no pain in prone position with knee flexed to 15°, no pain during slump test, < 10% asymmetry when in prone position with knee flexed to 15°, < 10% asymmetry during active knee extension test, and < 5° asymmetry in the modified Thomas Test.

The goals of the functional phase (Phase III) included to (i) increase the optimum length of the hamstrings, (ii) decrease leg asymmetries in optimum length, (iii) decrease leg asymmetries in concentric hip extension, (iv) decrease leg asymmetries in horizontal force production during running, and (v) improve torsional capabilities. The functional phase comprised daily exercises, with three sessions per week at the clinic of the PI (every other day) and the remaining sessions at the club or at home. The exercises comprised the following: core stability and lumbopelvic control, flexibility and neural mobilization, hamstring and gluteal strength, plyometric training, and running technique. During the functional phase, the running sessions consisted of two sets of ten minutes at moderate to high intensity (individually rated by the patient). Suspension of running sessions was permitted in the event of discomfort or pain.

The criteria for return to sport were (according to [27]): absence of pain on palpation, absence of pain during flexibility testing (active knee extension test and passive straight leg raise test), absence of pain during strength testing (isometric force test), absence of pain during and after functional testing (repeated sprint ability test and single leg bridge), similar hamstring flexibility, psychological readiness / athlete confidence, and clearance by the medical staff. The quantity and quality of the supervised rehabilitation sessions at home or the sports club were documented.

Patients in the rESWT group received the specific rehabilitation program as outlined above, and rESWT as follows: up to nine rESWT sessions; three sessions per week (interval between sessions: two or three days); rESWT device: Swiss DolorClast (Electro Medical Systems, Nyon Switzerland), EvoBlue handpiece, 15 mm applicator; 2500 radial extracorporeal shock waves (rESWs) per session, with energy flux density (EFD) between 0.12 and 0.16 mJ/mm^2^ (achieved by operating the Swiss DolorClast at air pressure between three and four bar), depending on what the patient tolerated; 15 rESWs per second, resulting in treatment time between three and five minutes per session; application of rESWs in prone position, with the patient lying on an examination table; exact location of the application of rESWs determined by clinical and ultrasonographic examinations; treatment of both the side of injury and the entire affected muscle (from distal to proximal in order to relax the affected muscle); application of rESWs in sagittal (dorsal – ventral) direction; and no use of local anesthesia.

Patients in the sham rESWT group received the specific rehabilitation program as outlined above, and sham rESWT as outlined above for rESWT, with a specially designed sham EvoBlue handpiece that looked and sounded like the EvoBlue handpiece of the Swiss DolorClast, but did not generate rESWs. This was achieved by blocking the projectile (“2” in Figure 1) shortly before it strokes the metal applicator (“4” in Figure 1). The sham EvoBlue handpiece did not emit any rESW energy.

**Figure 1.**
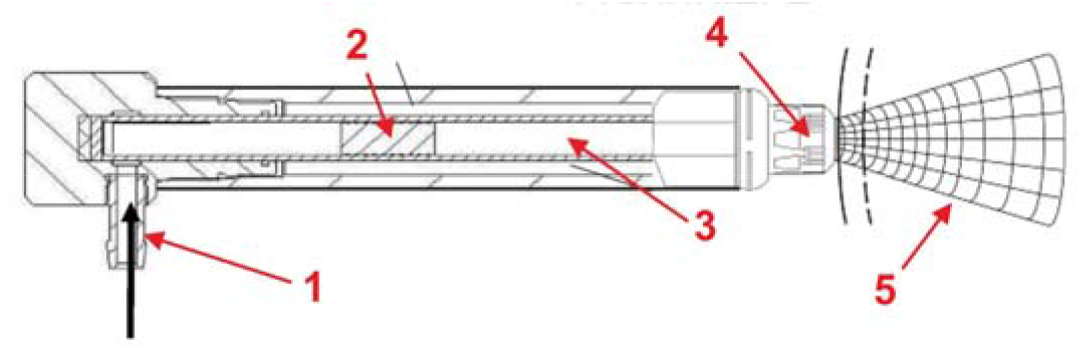
Working principle of the handpiece of a pneumatic, radial extracorporeal shock wave therapy (rESWT) device (modified from [9]). Compressed air (1) is used to fire a projectile (2) within a guiding tube (3) on a metal applicator (4) that is placed on the patient’s skin. The projectile generates stress waves in the metal applicator that transmit pressure waves (radial extracorporeal shock waves) into tissue (5).

### Outcome Measures

The primary outcome was the individual time (days) to return to sport with the criteria outlined above (according to [27]).

Secondary outcomes were individual patient’s satisfaction at six months post-inclusion into this trial (using a scale ranging from 0 (maximum dissatisfaction) to 10 (maximum satisfaction), and presence or absence of re-injury during a time period of six months post-inclusion into this trial (defined as sudden, sharp pain in the posterior aspect of the thigh that was initially injured, accompanied by the same objective criteria initially used for the diagnosis of acute HMC injury type 3b).

### Statistical Analysis

Missing data imputation was performed in three steps: (i) Little’s MCAR test was used to test the hypothesis that missing data was *missing completely at random* (MCAR); (ii) linear regression analysis was performed to test the hypothesis of a negative correlation between the durations of Phase II and Phase III of the RP; and (iii) missing data imputation was performed using the *Expectation Maximization* (EM) Technique, with a maximum of 50 iterations.

The primary outcome (individual time to return to sport) and the secondary outcome “patient’s satisfaction at six months post-inclusion into this trial” each returned a single data point for each patient. Assessing normality of these data was performed using the D’Agostino & Pearson test before deciding whether differences between groups were tested using the parametric Student’s t test or the nonparametric Mann-Whithney test.

The secondary outcome “presence or absence of re-injury during a time period of six months post-inclusion into this trial” returned a single data point (“yes”, “no” or “unknown” in case of missing data) for each patient. Comparison between groups was performed using the Chi-square test.

The probability value of less than 0.05 (p-value < 0.05) was considered statistically significant [28]. Calculation of Little’s MCAR test and the EM technique were performed using IBM SPSS Statistics (version 28.0.0.0; IBM, Armonk, NY, USA); all other calculations were performed using GraphPad Prism (version 10.2.3 for Windows, GraphPad Software, San Diego, CA, USA).

## RESULTS

### Compliance with Trial Method

This trial was carried out as described in the published protocol [18]. The only deviation from the protocol was that an experienced investigator (PS) who served as team physician of a German Bundesliga soccer club for many years re-examined all ultrasonography images to confirm the ultrasonographic diagnosis of acute HMC injury type 3b. For logistical reasons, however, this was only possible after the trial was completed. In 4 of 40 (10%) of the patients the ultrasonographic diagnosis of acute HMC injury type 3b could not be confirmed as the size of the lesion was smaller than 5 mm (resulting in the diagnosis of acute HMC injury type 3a). These patients were excluded from the analysis and were not replaced in the randomization scheme, resulting in a modified Intent-to-Treat (mITT) population of n=18+18=36.

### Baseline clinical characteristics

Table 1 shows the clinical characteristics of the patients in the mITT population at baseline (additional information is provided in Supplementary Information). 81% were male and the median / mean age was 26 years / 25.9 ± 4.9 (mean ± SD) years (range, 18 – 34 years). 47.2% of the patients played soccer, 27.8% hockey and 25% rugby. The median / mean length of time the sport was practiced was 11 years / 12.9 ± 4.3 years (range, 6 – 22 years), and the median / mean number of hours of sport practiced per week was 9 hours / 9.8 ± 1.3 hours (range, 8 – 14 hours). 69.4% of the injuries occurred during competition and 30.6% during training. 69.4% of these injuries affected the long head of the biceps muscle, 25.0% the semimembranosus muscle and 5.6% the semitendinosus muscle. According to a new classification of acute muscle strain injuries developed by one of us (NM) (outlined in [29]) 11.1% of the injuries occurred at the proximal musculotendinous junction and 88.9% within the affected muscle. Among the latter, 18.8% were located in the proximal part of the muscle, 50.0% in the middle of the muscle and 31.2% in the distal part of the muscle. The most frequently observed site of lesion was *myotendinous in the middle part of the muscle* (33.3% of all lesions), followed by *myofascial/perifascial in the distal part of the muscle* (16.7% of all lesions) and *myotendinous in the proximal part of the muscle* (13.9% of all lesions). The median / mean size of the lesion determined by ultrasonography during initial diagnosis was 7 mm / 8.1 ± 2.5 mm, and 9 mm / 9.5 ± 2.5 mm determined during re-examination.

**Table 1.** Baseline clinical characteristics of the study patients.

| Characteristic | rESWT + RP (n=18) | Sham rESWT + RP (n=18) |
| --- | --- | --- |
| Age [yr] (min / median / max / mean / SD) | 18 / 24.5 / 34 / 25 / 4.9 | 18 / 26 / 34 / 26.7 / 4.8 |
| Gender [n] (male / female / non-binary) | 14 / 4 / 0 | 15 / 3 / 0 |
| Body height [cm] (min / median / max / mean / SD) | 151 / 178 / 194 / 173.7 / 10.9 | 160 / 179.5 / 187 / 176.9 / 7.4 |
| Body weight [kg] (min / median / max / mean / SD) | 45 / 72 / 95 / 69.8 / 14.0 | 49 / 75.5 / 102 / 75.1 / 12.4 |
| BMI [kg/m<sup>2</sup>] (min / median / max / mean / SD) | 19.1 / 23.0 / 27.5 / 22.8 / 2.3 | 19.1 / 23.4 / 29.2 / 23.8 / 2.3 |
| Discipline [n] (soccer / hockey / rugby) | 9 / 5 / 4 | 8 / 5 / 5 |
| Years in sport (min / median / max / mean / SD) | 6 / 10 / 22 / 12.4 / 4.4 | 7 / 13.5 / 20 / 13.4 / 4.2 |
| Number of hours of sport practiced per week (min / median / max / mean / SD) | 8 / 9 / 14 / 9.7 / 1.4 | 8 / 9.5 / 12 / 9.8 / 1.3 |
| Symptoms 72 hours before enrollment (no / yes) | 14 / 4 | 14 / 4 |
| Time of event (match / training) | 13 / 5 | 12 / 6 |
| Activity at the moment of injury (running / stretching) | 14 / 4 | 14 / 4 |
| Side (left / right) | 5 / 13 | 13 / 5 |
| Injured muscle (long head of biceps / semimembranosus / semitendinosus) | 13 / 3 / 2 | 12 / 6 / 0 |
| Classification of the site of lesion [32]<sup>a</sup><br>(1 / 2AcD / 2Ad / 2Bb / 2Bcd / 2Bd / 2Be / 2Cc / 2Cd) | 2 / 1 / 3 / 0 / 1 / 7 / 0 / 2 / 2 | 2 / 0 / 2 / 1 / 1 / 5 / 1 / 4 / 2 |
| Size of the lesion determined during initial diagnosis [mm]<br>(min / median / max / mean / SD) | 5 / 7 / 15 / 7.9 / 2.6 | 5 / 7 / 12 / 8.3 / 2.4 |
| Size of the lesion determined during re-examination [mm]<br>(min / median / max / mean / SD) | 5 / 9.5 / 15 / 9.4 / 2.8 | 7 / 9 / 14 / 9.6 / 2.5 |

<sup>a</sup>, 1, proximal musculotendinous junction; 2, muscle; A, proximal; B, middle; C, distal; a, intramuscular; b, myofascial; c, myofascial/perifascial; d, myotendinous; e, combined.

### Flow of Patients through the Trial

The flow of patients through the trial is shown in Figure 2. One patient in the sham rESWT + RP group was lost during Phase 3 of the RP, and one patient in the rESWT + RP group was lost during the time period of six months post-inclusion into this trial. The missing data of these patients (patient in the sham rESWT + RP group: time to return to sport and satisfaction; patient in the rESWT + RP group: satisfaction) were imputed. Little’s MCAR test confirmed that missing data were *missing completely at random* (p = 0.163)

**Figure 2.**
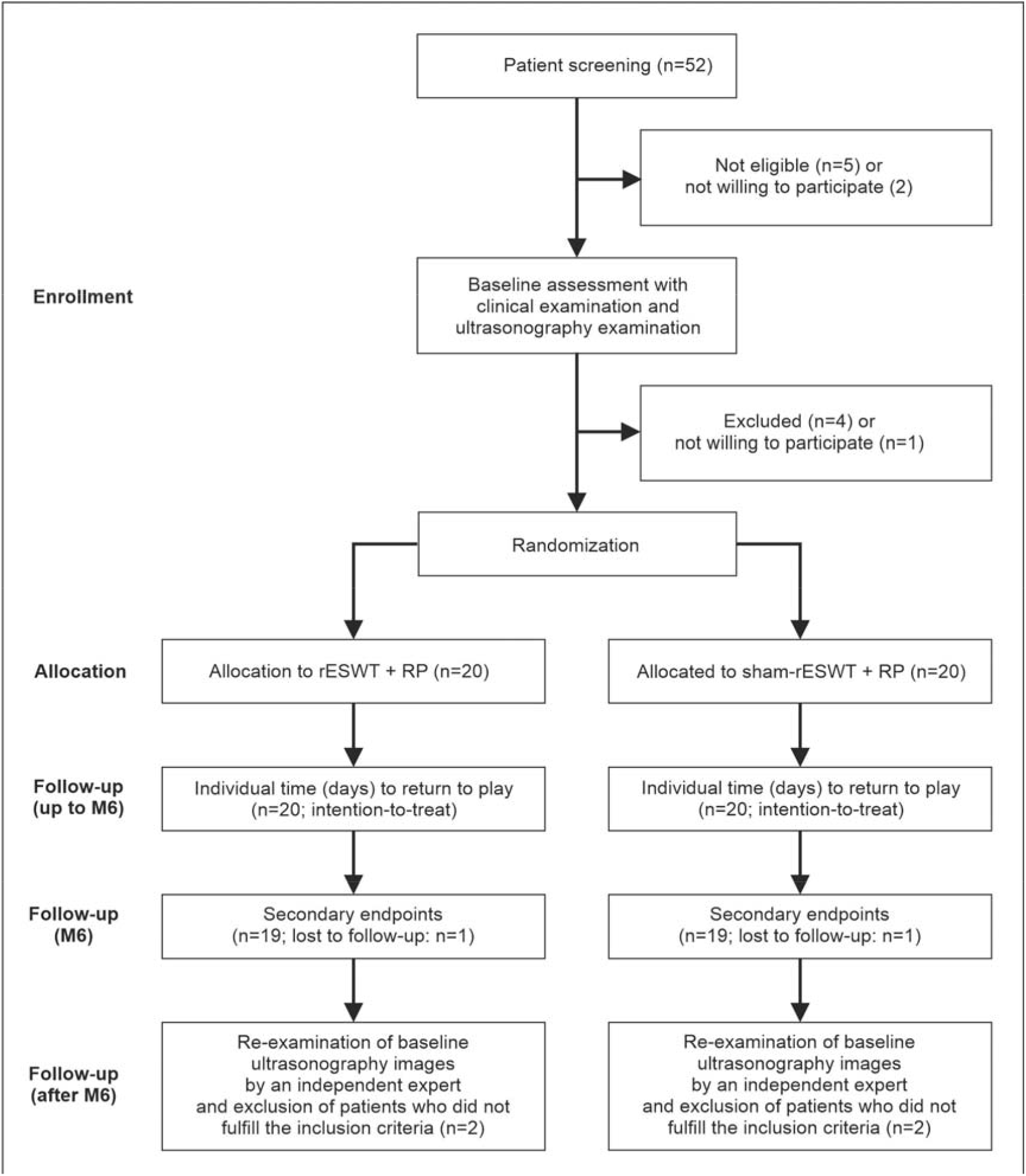
Flow of patients through the trial according to the CONSORT statement [30].

Figure 3 shows the time course of all patients in the mITT population during the trial. It was noticeable that some patients in the sham rESWT + RP group had a long Phase II and a short Phase III (e.g., patients 35 and 36), whereas other patients in this group had a short Phase II and a long Phase III (e.g., patients 21 and 30). The patients in the rESWT + RP group did not show this pattern. This finding raised the hypothesis of a negative correlation between the durations of Phase II and Phase III of the patients in the sham-rESWT + RP group, which was demonstrated using linear regression analysis (Fig. 4). Accordingly, the missing time to return to sport of patient 24 had to be imputed based on the combined durations of Phases II and III of the patients in the sham rESWT + RP group rather than on the duration of Phase III of these patients alone.

**Figure 3.**
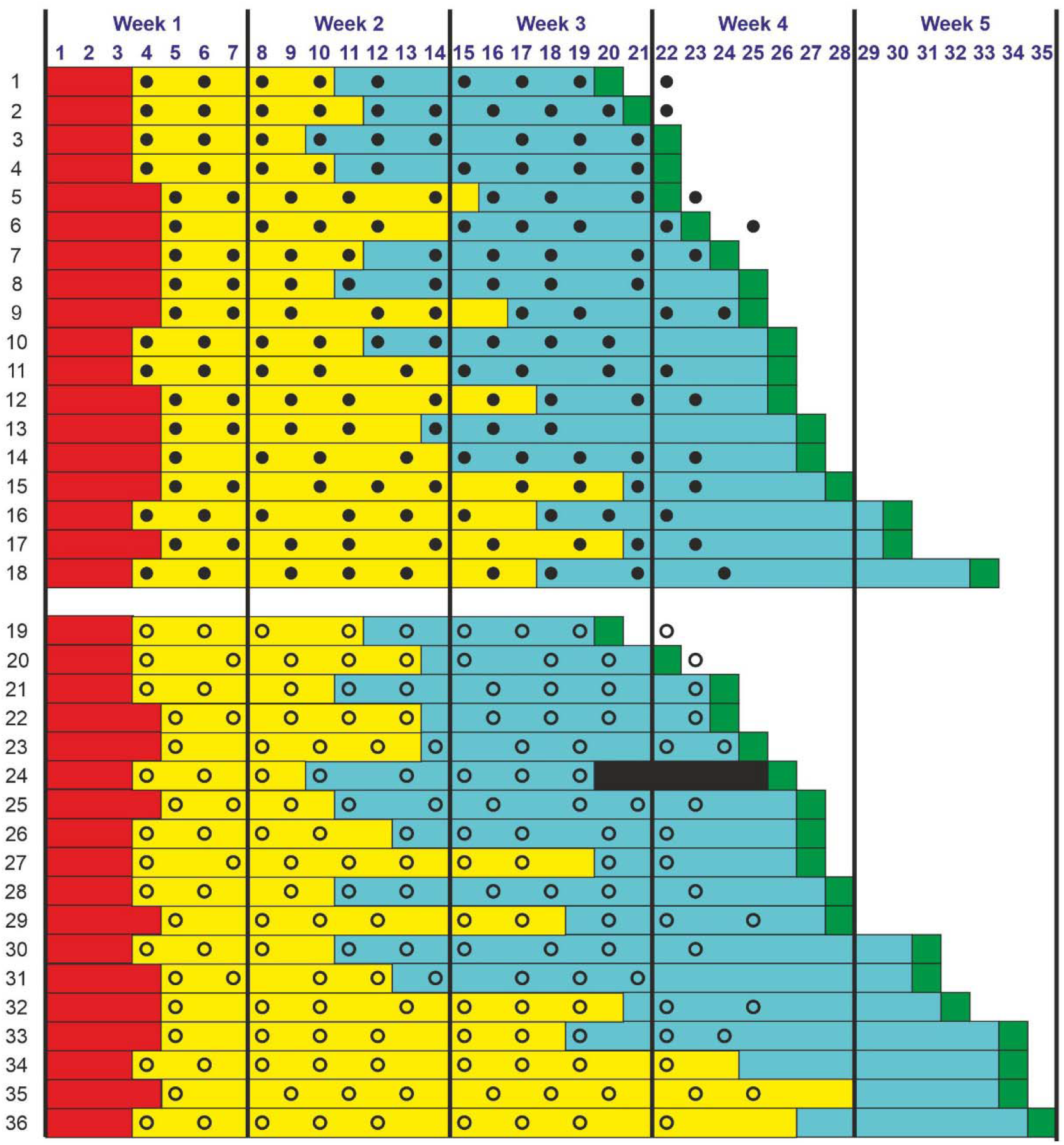
Individual time course of the patients in this trial. The horizontally arranged numbers are the days in the study (Day 1, randomization). The vertically arranged numbers represent the consecutive order of the patients according to the invidivual time to return to sport in the rESWT + RP group (patients 1-18) and the sham rESWT + RP group (patients 19-36) of the mITT population. For each patient the durations of Phase 1 (red), Phase 2 (yellow) and Phase 3 (blue) as well as the day of return to sport (green) are indicated, as well as all rESWT treatments (closed dots) or sham-rESWT treatments (open dots). The black rectangle in the line of patient 24 indicates the imputed missing value.

**Figure 4.**
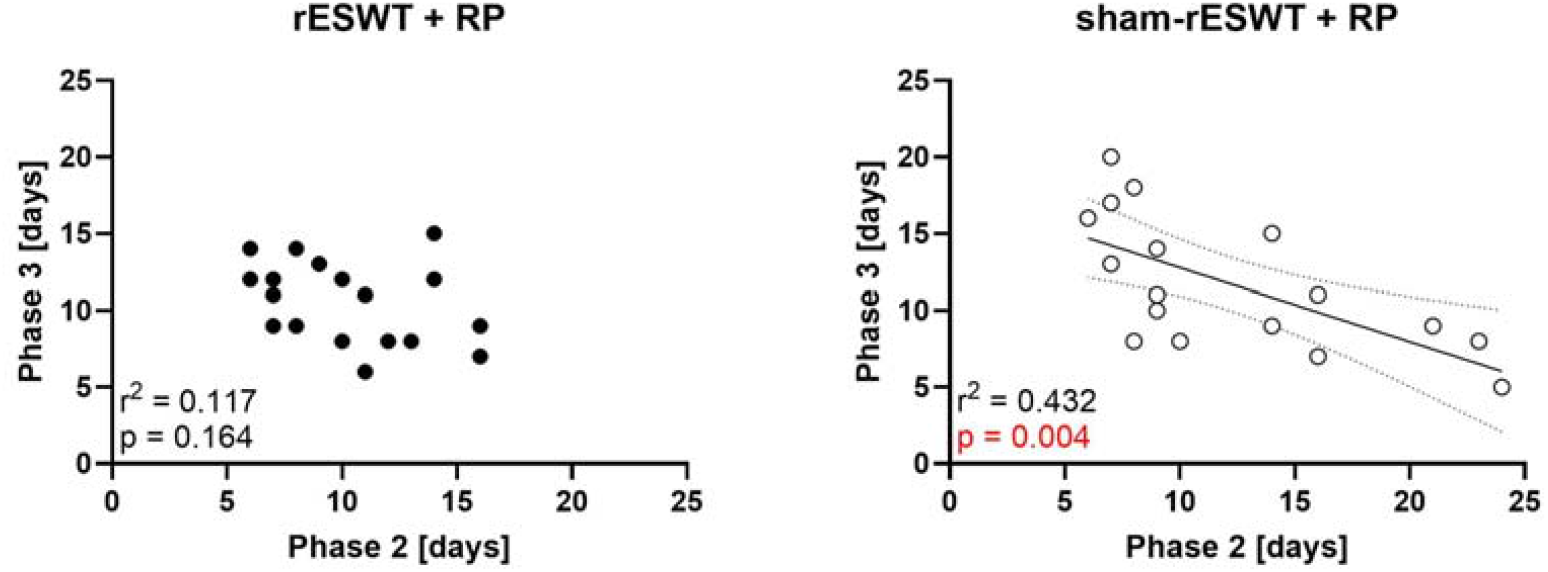
Duration of Phase 3 (functional phase) as a function of the duration of Phase 2 (subacute/regeneration phase) of all patients in this trial (except of patient 24 in the sham-rESWT + RP group who was lost during Phase III). The results of linear regression analysis are indicated.

### Time to Return to Sport

The time to return to sport data of both groups in the mITT population passed the D’Agostino & Pearson test for normal distribution (rESWT + RP group, p = 0.677; sham rESWT + RP group, p = 0.584). The median / mean time to return to sport of the patients in the rESWT + RP group was 25.5 days / 25.4 ± 3.5 days, and 27.5 / 28.3 ± 4.5 days of the patients in the sham rESWT + RP group (c.f. Table 2). This difference was statistically significant (unpaired, two-tailed Student’s t test; p=0.037).

**Table 2.** Results obtained for the modified Intent-to-Treat population (n=36).

| Outcome | rESWT + RP group<br>(n=18) | Sham rESWT + RP<br>group (n=18) | Sign. |
| --- | --- | --- | --- |
| Phase 1 [days] (min / median / max / mean / SD) | 3 / 4 / 4 / 3.6 / 0.5 | 3 / 3 / 4 / 3.4 / 0.5 | 0.740<sup>a</sup> |
| Phase 2 [days] (min / median / max / mean / SD) | 6 / 10 / 16 / 10.3 / 3.3 | 6 / 9 / 24 / 11.9 / 5.9 | 0.713<sup>a</sup> |
| Phase 3 [days] (min / median / max / mean / SD) | 6 / 11 / 15 / 10.6 / 2.6 | 5 / 11 / 20 / 11.7 / 4.3 | 0.560<sup>a,b</sup> |
| Time to return to sport [days] (min / median / max / mean / SD) | 20 / 25.5 / 33 / 25.4 / 3.5 | 20 / 27.5 / 35 / 28.3 / 4.5 | 0.037<sup>c</sup> |
| Satisfaction with treatment (min / median / max / mean / SD) | 7 / 9 / 10 / 9.0 / 1.1 | 7 / 8.5 / 10 / 8.4 / 1.2 | 0.145<sup>c</sup> |
| Re-injury within 6 months after inclusion (n) (no / yes / unknown) | 16 / 1 / 1 | 16 / 1 / 1 | >0.999<sup>d</sup> |

<sup>a</sup>, result of Mann-Whitney test (p value); <sup>b</sup>, without the missing data of patient 24 in the sham-rESWT + RP group; <sup>c</sup>, result of Student's t test (p value); <sup>d</sup>, result of Chi-square test (p value).

### Patient’s Satisfaction

The individual patient’s satisfaction data of both groups in the mITT population also passed the D’Agostino & Pearson test for normal distribution (rESWT + RP group, p = 0.276; sham rESWT + RP group, p = 0.115). The median / mean satisfaction of the patients in the rESWT + RP group was 9 / 9.0 ± 1.1, and 8.5 / 8.4 ± 1.2 days of the patients in the sham rESWT + RP group (c.f. Table 2). This difference was not statistically significant (unpaired, two-tailed Student’s t test; p=0.145).

### Re-injury Rate

16 of 18 (88.9%) patients in each group of the mITT population did not experience a re-injury during a period of six months post-inclusion into this trial. The latter was only experienced by one patient in each group. For one patient in each group this could not be determined because the corresponding patients were lost to follow-up (these missing data could not be imputed) (c.f. Table 2). The re-injury rate showed no statistically significant difference between the groups (Chi-square test; p > 0.999).

## DISCUSSION

The first key finding of this study was that the specific rehabilitation program applied in this study resulted in a median time to return to sport after acute HMC injury type 3b (sham rESWT + RP group: 27.5 days) that was shorter than corresponding time intervals reported by (i) Ekstrand et al. [31] for elite soccer players (moderate partial muscle tears (type 3b) of the posterior thigh muscles: 30 days; mean ± SD: 35.5 ± 19.5 days), (ii) Reurink et al. [8] for competitive and recreational athletes (median time to return to sport after injection of PRP as well after injection of isotonic saline as a placebo after hamstring lesion diagnosed on magnetic resonance imaging (MRI), defined as increased signal intensity on STIR and/or T2-weighted images, limited to one location in the muscle: 42 days) and (iii) Bayer et al. [22] for amateur athletes with acute injury of the thigh muscle (60% of the patients) or calf muscle (40% of the patients) confirmed on ultrasonography and MRI: 62.5 days with early therapy starting on day 2 post-injury or 83 days with delayed therapy starting on day 9 post-injury). Furthermore, the range of the times to return to sport of the patients in the sham rESWT + RP group of this study (20 – 35 days) was different from the corresponding range reported for type 3b muscle injuries in the guidelines of the Italian Society of Muscles, Ligaments and Tendons for muscle injuries (25 – 35 days) [6]. These differences could result from the selection of the treated patients, details of the respective treatment protocols, differences in the definition of return to sport, or complex interactions of the above-mentioned influencing factors. In any case, it seems attractive to use the specific rehabilitation program applied in this study as a control therapy in future studies on the treatment of muscle injuries

The second key finding of this study was that addition of rESWT to the specific rehabilitation program used in this study further reduced the median / mean time to return to sport by 2 days / 2.9 days or by 7.3% / 10.3%. At first glance, this further improvement, despite being statistically significant, appears to be unremarkable. On the other hand, this finding is of particular importance in professional sports, as a single day of lost activity by an athlete can cost several thousands of US dollars ($). For example, assuming that a professional athlete has a gross salary of $250,000, a reduction in the time to return to sport by three days would mean a saving of more than $2000 for the club, which justifies the costs of additional rESWT. The Global Sports Salaries Survey 2019 [32] may help to put these figures into context. According to this survey, the average annual salary of professional soccer players in 2019 was $1.4 million (m) in the French League 1, $2.1m in the German Bundesliga, $2.4m in the Italian Seria A, $2.7m in the Spanish La Liga and $4.2m in the British Premier League (calculated using an exchange rate of 1 British Pound = $1.326 on 31 December 2019). In none of the clubs in these leagues was the average annual salary less than $250,000 [32]. Furthermore, in the US National Basketball Association (NBA) the average annual salary of first-team players in 2019 varied between $7.1m (New York Nicks) and $10.0m (Portland Trail Blazers) [32].

Of note was the finding that further shortening the mean time to return to sport by rESWT did not cause any intercurrent events (including re-injury) in the investigated athletes. This is of particular importance as some authors warned against early mechanical stress on injured muscle tissue because of a potential risk of the formation of heterotopic ossification (HO) from the sense of myositis ossificans following extensive soft tissue trauma [3]. However, tissue stimulation by rESWT does not seem to exert this negative effect on injured muscle tissue. Rather, early application of rESWT on injured muscle tissue may block the specific molecular signals and receptors for the recruitment and/or differentiation of circulating cell populations contributing to heterotopic ossification, and thus may ultimately prevent the transition of preosseous tissue to definitive bone (c.f. [33]). Once HO is present in post-traumatic myositis ossificans, ESWT may attenuate the clinical symptoms but cannot be used to remove the HO itself [34, 35].

Several results from basic science studies and investigations of animal models suggest an explanation for the therapeutic success of rESWT in acute structural muscle injuries. One important mechanism in this regard is the direct stimulation of resident muscle cells to regenerate injured muscle tissue: in rats with experimentally induced, acute type 3 muscle tears application of rESWs (generated with the same rESWT device that was used in this study) resulted in a lower number of mononucleated cells and a higher number of newly formed muscle fibers at 4 days and 7 days post-treatment compared to treatment with diclofenac and no treatment (control) [14]. Furthermore, the rats treated with rESWT had higher levels of RNA expression of MyoD and Myosin, two markers characteristic of muscle regeneration [14]. Similar outcomes, including increased cell viability and the upregulation of the expression of the muscle specific genes Myf5, MyoD, Pax7 and NCAM, were observed when cultured human skeletal muscle cells were exposed with rESWs (also generated with the same rESWT device that was used in this study) [13].

Although pro-angiogenic gene expression is increased in skeletal muscle by ESWT, the actual density of blood vessels in the treated muscle tissue does not appear to increase after ESWT [14, 36]. Based on early reports of ESWT-induced increased expression of angiogenesis-related growth factors including eNOS (endothelial nitric oxide synthase), VEGF (vascular endothelial growth factor) and PCNA (proliferating cell nuclear antigen) [37], neoangiogenesis is a commonly cited effect of ESWT thought to be central to its mechanism of action and has been demonstrated in various tissues including the tendon-bone junction (focused ESWT; [38]) and the skin (rESWT; [39]), but does not appear to be involved in muscular regeneration.

Furthermore, rESWT has additional mechanisms of action that can promote muscular regeneration. For example, following an injury, the damaged muscle frequently develops excessive tone, which hinders regeneration and is occasionally addressed by infiltrating local anesthetics [3]. However, intramuscular injections of local anesthetics regularly result in reversible myonecrosis, with muscular regeneration within 3 to 4 weeks [40]. Reduction in muscle tone by rESWs without compromising muscle function is used in rESWT for spasticity [41-43]. Furthermore, Kenmoku et al. [44] demonstrated a dose-dependent reduction in the mean compound muscle action potential (CMAP) by exposing rat muscles in vivo with rESWs (in all of these studies [41-44] rESWs were generated with the same rESWT device that was used in the present study).

Regarding the treatment parameters of rESWT for acute HMC injury type 3b (i.e., the number of treatment sessions, number of treatment sessions per week, number of rESWs per treatment session, EFD and frequency of the rESWs), the current study protocol is a good starting point for orientation. However, fixed parameters are not as helpful in the practical application of rESWT as they are in the scientific setting. In this regard Morgan et al. [15] provided a retrospective case series of rESWT for acute muscle injuries type 1a, 2b and 3a in professional soccer players, using a highly individualized rESWT protocol that can also be applied to the treatment of acute HMC injury type 3b. Specifically, the EFD of the applied rESWs was adjusted individually in [15], with the athlete experiencing some discomfort but no pain during the treatment. The number of rESWs per treatment session was also individually adjusted between 6,000 and 12,000 rESWs, and with a few exceptions rESWT was performed daily or on every second day. Based on the individual patient’s response to rESWT, differences in the severity of muscular injury and several other influencing factors that vary between patients, future treatment and study protocols should also incorporate and implement individualized parameters.

### Relevance to Clinical Practice

After acute HMC injury type 3b the specific rehabilitation program applied in this study results in shorter mean / median times to return to sport than other rehabilitation programs reported in the literature [6, 8, 22, 31]. Furthermore, in situations where after acute HMC injury type 3b every day of lost activity counts, addition of rESWT to the specific rehabilitation program further reduces the time to return to sport without increased risk of re-injury or developing heterotopic ossification.

### Methodological Considerations

This study has three main limitations. First, establishing ultimate evidence that the combination of rESWT and the specific rehabilitation program applied in this study is superior to the rehabilitation program alone with respect to reduced times to return to sport will require to investigate a larger cohort of injured athletes. Specifically, based on the times to return to sport reported in this study (rESWT + RP group: 25.4 ± 3.5 (mean ± SD) days; sham rESWT + RP group: 28.3 ± 4.5 days) one will have to enroll at least 2 × 31 = 62 (or 2 × 41 = 82) patients into a RCT in order to demonstrate superiority of rESWT + RP over sham rESWT + RP with a two-sided confidence interval of 95% and a power of 80% (or 90%) (calculations were performed with the software, Open Source Epidemiologic Statistics for Public Health [45]). The reason for the relatively small sample size calculated in the protocol of this study [18] was our (PS’ and CS’) experience in treating elite athletes suffering from HMC injury type 3b with rESWT, including a professional soccer player at a European top club (regularly playing in the UEFA Champions League and the FIFA World Cup) who incurred a HMC injury type 3b and returned to play (full 90-min competitive match) 35 days later, as well as the fact that in two studies published in the New England Journal of Medicine [8, 22] the cumulative probability of return of sport on day 35 after acute HMC injury type 3b was only respectively 20% [8] or 5% [22] after treatment with a rehabilitation program. However, it turned out that the specific rehabilitation program applied in this study resulted in shorter mean / median times to return to sport than the rehabilitation programs applied in [8, 22].

Second, diagnosis of acute HMC injury type 3b in this study was based on ultrasonography (US) rather than on MRI. The reason for this decision was the fact that in Argentina the costs of MRI scans for muscle injuries would simply be too high in relation to the athletes’ salaries (with a few exceptions among professional athletes). On the other hand, both US and MRI are effective to identify hamstring strains and tendinopathy (e.g., [6, 46-48]), and both US and MRI provide detailed information about the HMC with respect to localization and characterization of injury [46].

Third, 10% of the patients enrolled in this RCT were retrospectively excluded because the ultrasonographic diagnosis of acute HMC injury type 3b could not be confirmed by an experienced investigator (PS) who served as team physician of a German Bundesliga soccer club for many years. It would have been advantageous to perform this independent assessment of ultrasonography scans before including the patients in the study, but unfortunately this was not possible for logistical reasons. On the other hand, including the results of all 40 patients enrolled in this study (provided in Supplementary Information) in the final analysis would have resulted in min / median / max / mean ± SD times to return to sport of 20 days / 26 days / 33 days / 25.8 ± 3.5 days of the patients in the rESWT + RP group, and 20 days / 27.5 days / 35 days / 28.2 ± 4.7 days of the patients in the sham rESWT + RP group, with a p value of 0.069 (unpaired, two-tailed Student’s t test). This, however, would be the result of a different study on patients with acute HCM injuries type 3a and 3b, which was not the purpose of this study. Accordingly, reporting the results of the mITT population of this study appears to be justified.

## CONCLUSIONS

In rehabilitation of athletes with acute HMC injury type 3b where every day to return to sport counts, the combination of rESWT and the specific rehabilitation program applied in this study is more effective than the specific rehabilitation program alone, and the latter is superior to other rehabilitation programs described in the literature.

## STATEMENTS AND DECLARATIONS

### Ethics approval

The Ethics Board of the Universidad Abierta Interamericana, Buenos Aires, Argentina (Nr. 0-1027).

### Data availability

The raw data are provided in Supplementary Information.

### Author contributions

Conception and design of the study: JC, SS, NW, AR, PS, CS. Data acquisition, analysis or interpretation of data: JC, SS, NW, AR, TW, PS, AM, JF, NM, CS. Drafting the manuscript: JC, TW, CS. All authors critically reviewed the manuscript for important intellectual content and approved the final version.

## Supplementary Information

This Supplementary Information contains additional information about the following variables of the patients in this trial.

| Variable | Description |
| --- | --- |
| V1 | Treatment in addition to the specific rehabilitation program |
| V2 | Activity at the moment of injury (running / stretching) |
| V3 | Injured muscle (long head of biceps / semimembranosus / semitendinosus) |
| V4 | Classification of the site of lesion<sup>1</sup> |
| V5 | Size of the lesion determined during initial diagnosis [mm] |
| V6 | Size of the lesion determined by retrospective re-assessment [mm] |
| V7 | Duration of Phase I [days] |
| V8 | Duration of Phase II [days] |
| V9 | Duration of Phase III [days] |
| V10 | Time to return to sport [days] |

<sup>1</sup> Classification according to Chan O, Del Buono A, Best TM, Maffulli N. Acute muscle strain injuries: a proposed new classification system. *Knee Surg Sports Traumatol Arthrosc.* 2012;20:2356-2362.

| No<sup>2</sup> | V1 | V2 | V3 | V4<sup>3</sup> | V5 | V6 | V7 | V8 | V9 | V10 |
| --- | --- | --- | --- | --- | --- | --- | --- | --- | --- | --- |
| 1 | rESWT | running | long head of biceps | 2,B,d | 6 | 7 | 3 | 7 | 9 | 20 |
| 2 | rESWT | stretching | semimembranosus | 1,d/c | 15 | 15 | 3 | 8 | 9 | 21 |
| 3 | rESWT | running | semimembranosus | 2,B,d | 6 | 6 | 3 | 6 | 12 | 22 |
| 4 | rESWT | running | long head of biceps | 2,C,d | 5 | 5 | 3 | 7 | 11 | 22 |
| 5 | rESWT | running | long head of biceps | 2,B,d | 7 | 10 | 4 | 11 | 6 | 22 |
| 6 | rESWT | running | long head of biceps | 2,C,d | 7 | 10 | 4 | 10 | 8 | 23 |
| 7 | rESWT | stretching | long head of biceps | 2,B,d | 5 | 6 | 4 | 7 | 12 | 24 |
| 8 | rESWT | running | long head of biceps | 2,A,d | 6 | 7 | 4 | 6 | 14 | 25 |
| 9 | rESWT | running | long head of biceps | 2,B,d | 7 | 9 | 4 | 12 | 8 | 25 |
| 10 | rESWT | running | long head of biceps | 2,C,b | 6 | 9 | 3 | 8 | 14 | 26 |
| 11 | rESWT | running | long head of biceps | 2,A,d | 8 | 8 | 3 | 11 | 11 | 26 |
| 12 | rESWT | running | long head of biceps | 2,C,c | 10 | 11 | 4 | 13 | 8 | 26 |
| 13 | rESWT | running | long head of biceps | 2,B,d | 7 | 13 | 4 | 9 | 13 | 27 |
| 14 | rESWT | stretching | semimembranosus | 2,B,d/c | 7 | 9 | 4 | 10 | 12 | 27 |
| 15 | rESWT | stretching | semitendinosus | 1,c/d | 10 | 10 | 4 | 16 | 7 | 28 |
| 16 | rESWT | running | long head of biceps | 2,A,d | 12 | 15 | 3 | 14 | 12 | 30 |
| 17 | rESWT | running | long head of biceps | 2,B,d | 10 | 10 | 4 | 16 | 9 | 30 |
| 18 | rESWT | running | semitendinosus | 2,A,d/c | 9 | 10 | 3 | 14 | 15 | 33 |
| 19 | sham rESWT | running | semimembranosus | 2,A,d | 9 | 12 | 3 | 8 | 8 | 20 |
| 20 | sham rESWT | running | long head of biceps | 2,B,e | 9 | 12 | 3 | 10 | 8 | 22 |
| 21 | sham rESWT | running | semimembranosus | 2,B,d | 7 | 10 | 3 | 7 | 13 | 24 |
| 22 | sham rESWT | stretching | semimembranosus | 2,C,c | 7 | 7 | 4 | 9 | 10 | 24 |
| 23 | sham rESWT | running | long head of biceps | 2,C,c | 7 | 7 | 4 | 9 | 11 | 25 |
| 24 | sham rESWT | stretching | semimembranosus | 2,B,d | 12 | 12 | 3 | 6 | 16<sup>4</sup> | 26 |
| 25 | sham rESWT | running | long head of biceps | 2,B,d | 5 | 8 | 4 | 6 | 16 | 27 |
| 26 | sham rESWT | running | long head of biceps | 2,C,d | 6 | 8 | 3 | 9 | 14 | 27 |
| 27 | sham rESWT | running | long head of biceps | 2,B,d | 9 | 9 | 3 | 16 | 7 | 27 |
| 28 | sham rESWT | running | long head of biceps | 2,C,c | 7 | 9 | 3 | 7 | 17 | 28 |
| 29 | sham rESWT | running | long head of biceps | 1,d | 12 | 12 | 4 | 14 | 9 | 28 |
| 30 | sham rESWT | running | long head of biceps | 2,B,d | 7 | 8 | 3 | 7 | 20 | 31 |
| 31 | sham rESWT | running | long head of biceps | 2,A,d | 12 | 12 | 4 | 8 | 18 | 31 |
| 32 | sham rESWT | running | long head of biceps | 1,d/c | 12 | 14 | 4 | 16 | 11 | 32 |
| 33 | sham rESWT | stretching | semimembranosus | 2,B,d/c | 10 | 10 | 4 | 14 | 15 | 34 |
| 34 | sham rESWT | running | long head of biceps | 2,C,c | 6 | 8 | 3 | 21 | 9 | 34 |
| 35 | sham rESWT | stretching | long head of biceps | 2,C,d | 5 | 7 | 4 | 24 | 5 | 34 |
| 36 | sham rESWT | running | semimembranosus | 2,B,b | 7 | 7 | 3 | 23 | 8 | 35 |
| <sup>5</sup> | rESWT | running | long head of biceps | 2,B,b | 7 | <5 | 4 | 16 | 7 | 28 |
| <sup>5</sup> | rESWT | running | semitendinosus | 2,B,d | 7 | <5 | 3 | 14 | 12 | 30 |
| <sup>5</sup> | sham rESWT | running | long head of biceps | 2,A,d | 5 | <5 | 3 | 24 | 6 | 34 |
| <sup>5</sup> | sham rESWT | running | semimembranosus | 2,B,d | 6 | <5 | 4 | 13 | 3 | 21 |

<sup>2</sup> These numbers represent the consecutive order of the patients according to the individual time to return to sport in the rESWT + RP group (patients 1-18) and the sham rESWT + RP group (patients 19-36) in the mITT population. They are identical to the corresponding numbers in Figure 3 in the manuscript.
<sup>3</sup> Abbreviations: 1, proximal musculo-tendinous junction; 2, muscle; A, proximal; B, middle; C, distal; a, intramuscular; b, myofascial; c, myofascial/perifascial; d, myotendinous; e, combined.
<sup>4</sup> Missing data imputed using the *Expectation Maximization Technique*.
<sup>5</sup> These patients were retrospectively excluded because the ultrasonographic diagnosis of acute HMC injury type 3b could not be confirmed by an experienced investigator (PS) who served as team physician of a German Bundesliga soccer club for many years.

